# Assessment of Experiences of Preventive Measures Practice including Vaccination History and Health Education among Umrah Pilgrims in Saudi Arabia, 1440H-2019

**DOI:** 10.1101/2020.06.09.20126581

**Authors:** Mansour Tobaiqy, Sami S. Almudarra, Manal M.Shams, Samar A. Amer, Mohamed F.Alcattan, Ahmed H.Alhasan

## Abstract

**Background:** Annually, approximately 10 million Umrah pilgrims travel to the Kingdom of Saudi Arabia for Umrah from more than 180 countries. This event presents major challenges for the Kingdom’s public health sector, which strives to decrease the burden of infectious diseases and to adequately control its spread.

**Aims of the study:** The aims of the study were to assess the experiences of preventive measures practice, including vaccination history and health education, among Umrah pilgrims in Saudi Arabia.

**Methods:** A cross sectional survey administered to a randomly selected group of pilgrims by the research team members from February to the end of April 2019 at the departure lounge at King Abdul Aziz International airport, Jeddah city. The questionnaire was comprised of questions on the following factors: sociodemographic information, level of education, history of vaccinations and chronic illnesses, whether the pilgrim has received any health education and orientation prior to coming Saudi Arabia or on their arrival, and their experiences with preventive medicine.

**Results:** Pilgrims (n=1012) of 48 nationalities completed the survey and were reported in this study. Chronic diseases (n=230) were reported among pilgrims, with hypertension being the most reported morbidity (n=124, 53.9%). The majority of pilgrims had taken immunization prior to travel to Saudi Arabia, and the most commonly reported immunizations were meningitis (n=567, 56%), influenza (n=460, 45.5%), and Hepatitis B virus vaccinations (n=324, 32%); however, 223(22%) had not received any vaccinations prior to travel, including meningitis vaccine, which is mandatory in Saudi Arabia. 305 pilgrims (30.1%) had reported never using face masks in crowded areas; however, 63.2% reported lack of availability of these masks.

The majority of participants had received health education on preventive measures, including hygienic aspects (n=799, 78.9%) mostly in their home countries (n=450, 56.3%). A positive association was found between receiving health education and practicing of preventive measures, such as wearing masks in crowded areas (P= 0.04) and other health practice scores (P= 0.02).

**Conclusion:** Although the experiences of the preventive measures among pilgrims in terms of health education, vaccinations, and hygienic practices were overall positive, this study identified several issues with the following preventive measures: immunizations particularly meningitis vaccine and using face masks in crowded areas.

Further studies are required to develop a health education module to promote comprehensive preventive measures for pilgrims.

## Introduction

Around 10 million pilgrims annually travel to the Kingdom of Saudi Arabia (KSA) for Umrah (minor pilgrimage to Makkah) from more than 180 countries (1). According to The Ministry of Hajj and Umrah in Saudi Arabia, the government has issued 7,584,424 Umrah visas since May 30th, 2019 (25 Ramadan 1440 H), with the majority coming via air, land, or sea and visiting the holy cities of Makkah and Madinah (1).

On February 27^th^, 2020, the Foreign Ministry of Saudi Arabia took a decisive and proactive preventative measure to prevent the arrival and subsequent spreading of the novel coronavirus (COVID-19) to the Kingdom by suspending the entry of pilgrims to the holy cities and the entire kingdom (2).

Umrah is a spiritual practice that is not obligatory and has no fixed date or time to be performed (3), and similarly to the Hajj and other mass gatherings, the Umrah entails an increased health challenges (4) including the risk of spreading infection. Respiratory tract infections, overcrowding accidents, pollution, and many other medical and health issues are encountered by the hosting country and require special attention and measurable responses to mitigate these risks (4, 5).

Lack of proper planning and preventive measures, this religious activity can overwhelm the health system of the host country and impact global health preparedness because the majority of Umrah pilgrim come from different countries of return and origin (5, 6).

The Umrah performer (pilgrim), called *hajji* in the Arabic language, must be received and accompanied by an authorized company by the Ministry of Hajj called “Al Hamla,” which provides Hajj and Umrah services including catering, transportation, travel arrangements, accommodations, and first aid health services for every pilgrim who paid for their services (1, 6).

Before travelling to Saudi Arabia for Umrah, pilgrims should seek medical advice about potential health risks and provide vaccination certificates to be inspected at the entry port by Saudi authorities (3). The Saudi Arabia Ministry of Health (MOH) has published a guide report for pilgrims on health issues and standard vaccination requirements, together with important control measures for the safety of the individuals and prevention of infectious disease outbreaks (7).

There is no doubt that providing health education to pilgrims on non-communicable and infectious diseases (Including the preventive measures and modes of infection transmission) is of great interest for the Hajj and health authorities in Saudi Arabia (1), it’s also critical for global public health and disease control; this is considered good practice to minimize the risks and improve the compliance with preventive control measures in several studies (8, 10).

During Umrah, pilgrims have a vital role in sustaining public health through practicing preventive measures such as personal hygiene, hand washing with water and soap or sanitizers, properly wearing face masks, and proper waste disposal in sanitary bins (5,7).

Face mask use is an affordable and effective method to control the exposure to pathogens and high-risk environments, to reduce the risk of transmittable infectious diseases including COVID-19 (11), and to protect from the inhalation of aerosols containing organic and inorganic particulates (12).

Following Saudi Arabia Vision 2030, which aims to build an outstanding health and economic services to serve the country visitors including Umrah and Hajj pilgrims, and in light of an initiative called “To care about Rahman Guests” introduced by the Ministry of Health Community Services in Jeddah in collaboration with voluntary group (Vision Team), and in view of the current global pandemic of COVID-19 that has affected millions of people worldwide, and the need for effective preventive measures to be adopted nationally to increase the preparedness for Umrah and Hajj during the suspension of restrictions, this study aims to assess the experiences of preventive measures practice, including vaccination history and health education, among Umrah pilgrims in Saudi Arabia.

## Methods

### Study design

A cross-sectional survey was conducted among Umrah pilgrims before departure from February to the end of April 2019 at King Abdul Aziz Airport-Islamic port in Jeddah, Saudi Arabia. Participation in the research was entirely anonymous and voluntary.

### Questionnaire development

The questionnaire comprised items on the following aspects; sociodemographic, level education, history of vaccinations and chronic illnesses, whether the pilgrim has received any health education and orientation prior to coming Saudi Arabia or upon their arrival, their experiences of the practice of preventive measures such as using umbrellas, hats, or sunscreen to protect from excessive heat and sun, use of hygienic tools and practices such as wearing facial masks and washing hands, and the availability and the sources of these tools. Pilgrims were also asked to rate the level of satisfaction around the health services they received and encountered by the Saudi Arabian ministry of health different facility and staff.

Item types included closed questions, where the tick list was originated from a combination of previous research on mass gathering and public health issues, with respondents permitted to tick more than one response in some questions.

The questionnaire was reviewed for face and content validity by members of research team and experts in public health in the Ministry of Health, Validity testing was followed by piloting with twenty Umrah pilgrims of different nationalities and spoken languages, and no changes were deemed appropriate post-piloting, therefore the piloted responses were included in the analysis dataset for reporting. The questionnaire was written in Arabic and was translated by the Al Hamla company’s supervisors to the pilgrim’s specific language to be easily understood. Finally, all responses were gathered by the research team and translated to English for analysis and reporting.

### Recruitment

Potential participants were approached opportunistically by a research team member from February until the end of April 2019 at the departure lounge at King Abdul Aziz International Airport in Jeddah city, and were asked if they have spare time to participate in a short survey concerning the preventive measures in Umrah practice. Those who agreed were administered the questionnaire by the 22 trained personnel who recorded the responses electronically into an automated dataset that excluded incomplete questionnaires. Participants had to be at least 18 years of age; there were no exclusion criteria.

### Analysis

Statistical analysis was undertaken using SPSS (SPSS Ic, Cart, NC version 22), The data was summarized and analyzed using suitable methods and tests. A T-test for independent samples was conducted to identify the relationship between receiving health education and the practice of preventive measures by pilgrims; p < 0.05, was considered statically significant.

### Ethics Approval

The study obtained permission from the Health Directorate Affairs in Jeddah Ministry of Health, reviewed by the College of Applied Medical Sciences Ethics & Scientific Committee at the University of Jeddah, and also received permission from the King Abdul Aziz Airport, Islamic Port in Jeddah. Authorization was given to all participant researchers and data collectors to obtain access to the airport and to interview pilgrims.

## Results

1,012 questionnaires were completed by the participants. 48 nationalities for the participants were reported in this study. The majority of pilgrims were from Egypt (n=188, 18.5%), followed by Sudan (n=113, 11.1%), Algeria (n=110, 10.6%), Iraq (n=79, 7.8%), Turkey (n=63, 6.2%), India (n=63, 6.2%), Morocco (n=54, 5.3%), Tunisia (n=48, 4.7%), and Indonesia (n=41, 4%).

### Sociodemographic characteristics

The majority of participants were males (n=656, 64.8%) and 35.2% were females (n=350), and the majority were above the age of 35 (n=342, 33.8% and married (n=773, 76.3). This is depicted in Table 1.

**Table 1.**
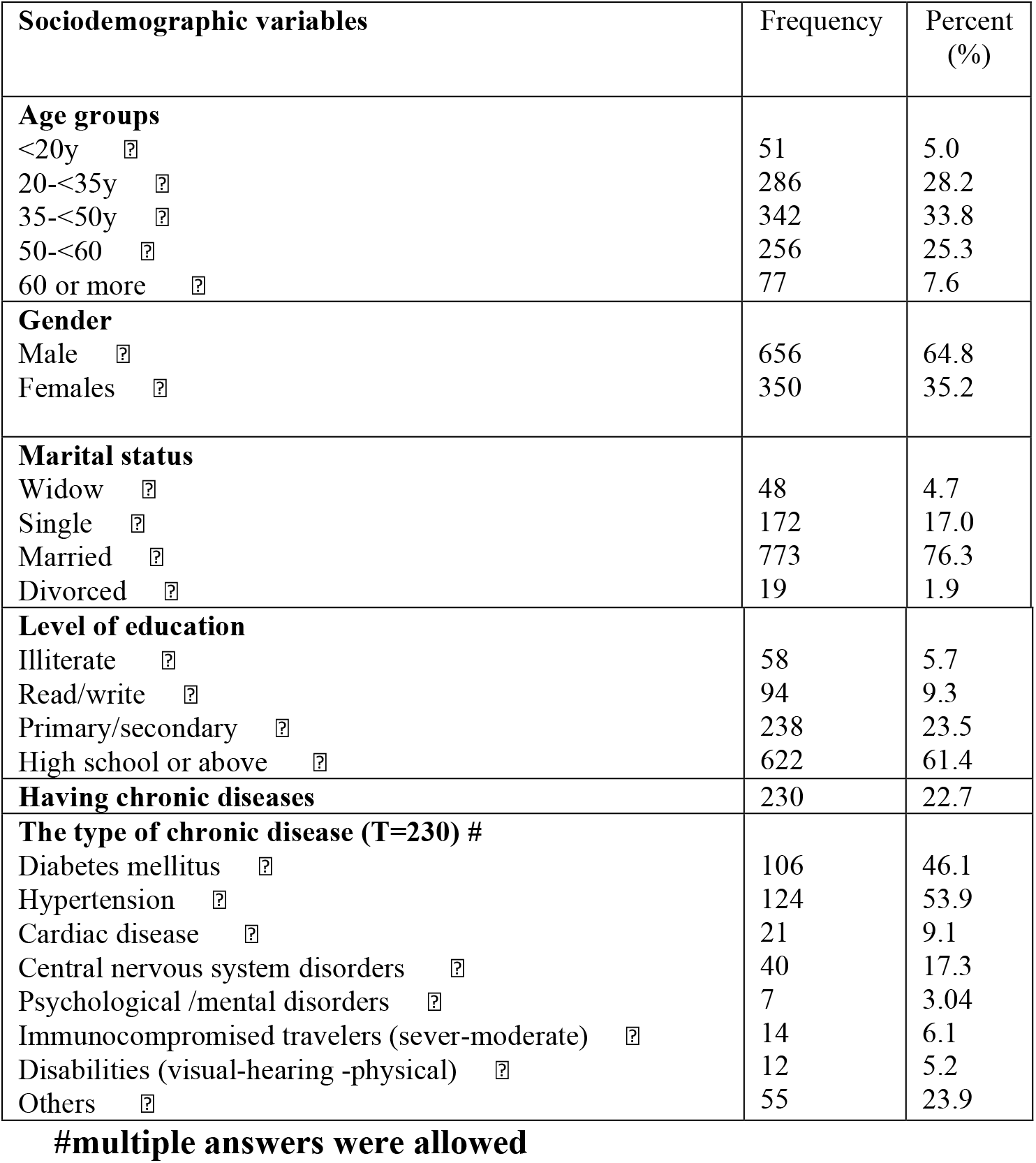
Responses to items on sociodemographic, level of education and chronic diseases of participated pilgrims (n=1012)

| **Sociodemographic variables** | **Frequency** | **Percent (%)** |
| --- | --- | --- |
| **Age groups** |  |  |
| <20y | 51 | 5.0 |
| 20-<35y | 286 | 28.2 |
| 35-<50y | 342 | 33.8 |
| 50-<60 | 256 | 25.3 |
| 60 or more | 77 | 7.6 |
| **Gender** |  |  |
| Male | 656 | 64.8 |
| Females | 350 | 35.2 |
| **Marital status** |  |  |
| Widow | 48 | 4.7 |
| Single | 172 | 17.0 |
| Married | 773 | 76.3 |
| Divorced | 19 | 1.9 |
| **Level of education** |  |  |
| Illiterate | 58 | 5.7 |
| Read/write | 94 | 9.3 |
| Primary/secondary | 238 | 23.5 |
| High school or above | 622 | 61.4 |
| **Having chronic diseases** | 230 | 22.7 |
| **The type of chronic disease (T=230) #** |  |  |
| Diabetes mellitus | 106 | 46.1 |
| Hypertension | 124 | 53.9 |
| Cardiac disease | 21 | 9.1 |
| Central nervous system disorders | 40 | 17.3 |
| Psychological /mental disorders | 7 | 3.04 |
| Immunocompromised travelers (sever-moderate) | 14 | 6.1 |
| Disabilities (visual-hearing -physical) | 12 | 5.2 |
| Others | 55 | 23.9 |

**#multiple answers were allowed**

### Level of education

Concerning the level of education, the majority had attended high school or above (n=622, 61.4%), and the minority were either illiterate (n=58, 5.7%) or only read and write (n=94, 9.3%). This data is provided in Table 1.

### Medical condition

Two hundred and thirty chronic diseases among pilgrims were reported, some had reported more than one morbidity. Hypertension (n=124, 53.9%) was the most reported chronic diseases, followed by diabetes mellitus (n=106, 46.1%) and central nervous system disorders (n=40, 17.3). This is data is provided in Table 1.

### Immunization history

223 pilgrims (22%) had not received any vaccination prior to travel; however, the majority had received meningitis vaccination (n=567, 56%), influenza vaccine (n=460, 45.5%), and Hepatitis B virus vaccine (HBV) (n=324, 32%) respectively (Table 2).

**Table 2.**
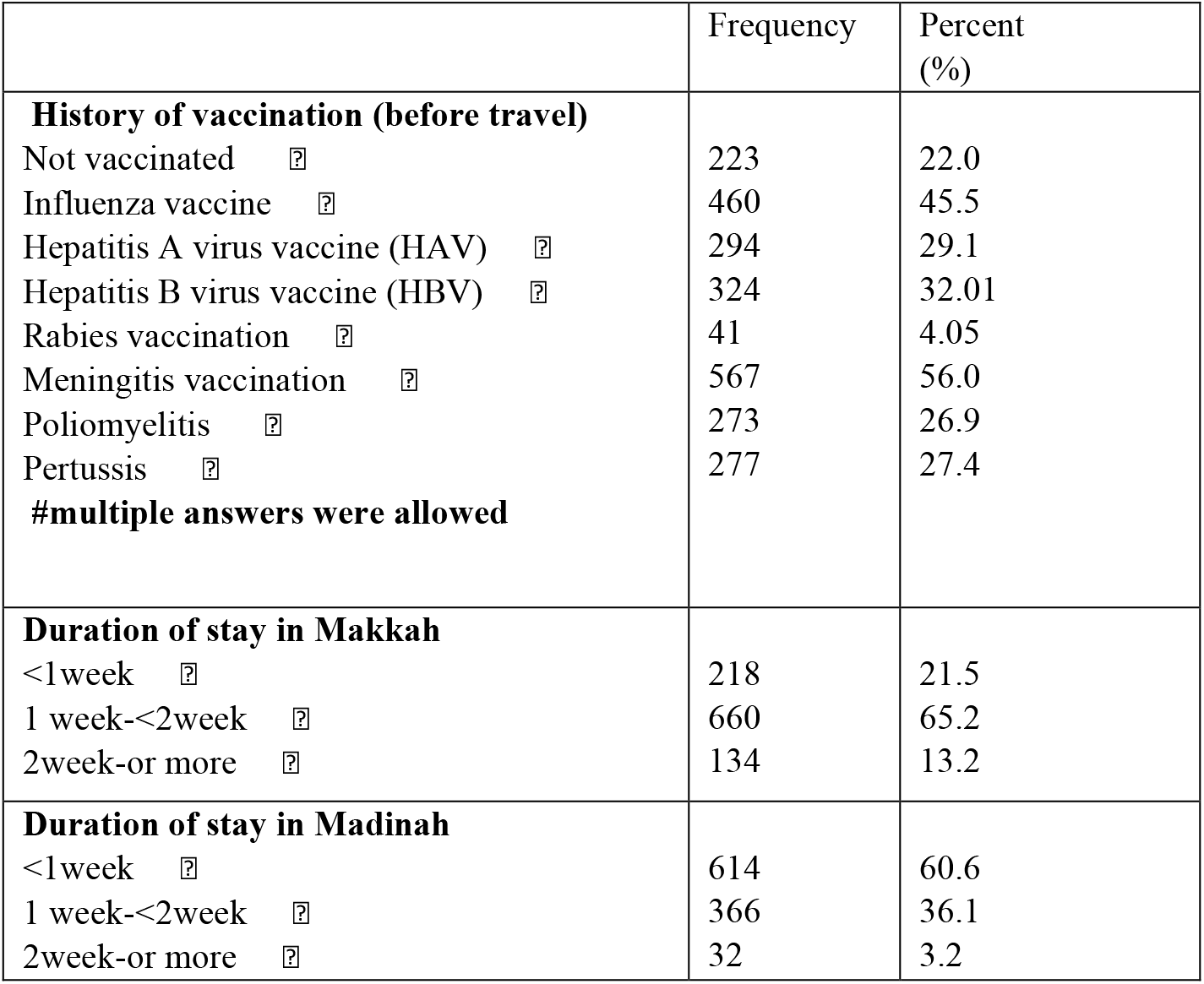
Responses to items on aspects of vaccination history and the average duration of stay in Makkah and Madinah (n=1012)

**Table 3.**
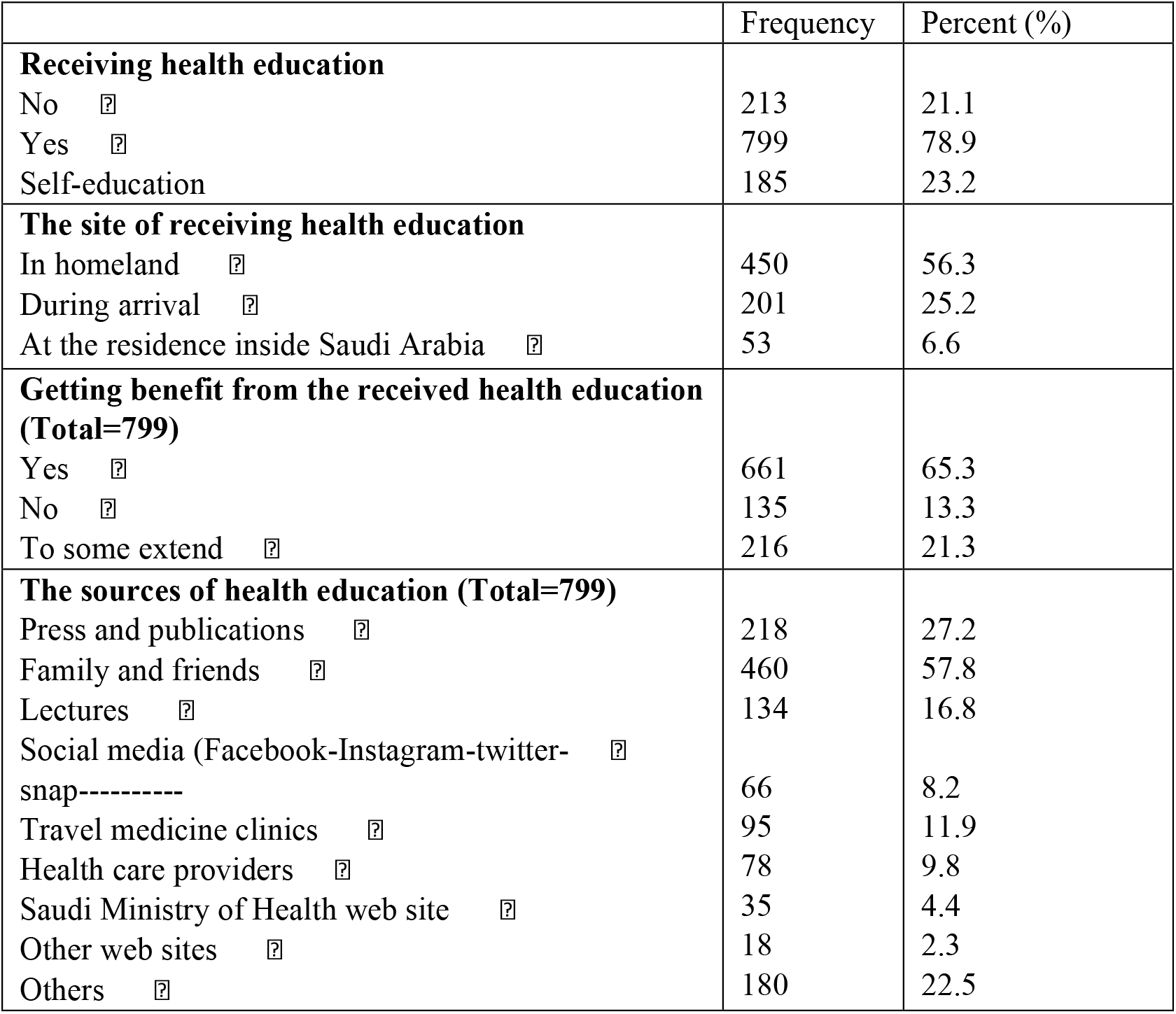
Responses to items on issues receiving health education (n=1012)

|  | Frequency | Percent (%) |
| --- | --- | --- |
| **Receiving health education** |  |  |
| No | 213 | 21.1 |
| Yes | 799 | 78.9 |
| Self-education | 185 | 23.2 |
| **The site of receiving health education** |  |  |
| In homeland | 450 | 56.3 |
| During arrival | 201 | 25.2 |
| At the residence inside Saudi Arabia | 53 | 6.6 |
| **Getting benefit from the received health education (Total=799)** |  |  |
| Yes | 661 | 65.3 |
| No | 135 | 13.3 |
| To some extend | 216 | 21.3 |
| **The sources of health education (Total=799)** |  |  |
| Press and publications | 218 | 27.2 |
| Family and friends | 460 | 57.8 |
| Lectures | 134 | 16.8 |
| Social media (Facebook-Instagram-twitter-snap-----) | 66 | 8.2 |
| Travel medicine clinics | 95 | 11.9 |
| Health care providers | 78 | 9.8 |
| Saudi Ministry of Health web site | 35 | 4.4 |
| Other web sites | 18 | 2.3 |
| Others | 180 | 22.5 |

### Duration of stay in Makkah and Madinah

The majority of participants stayed in Makkah for less than 2 weeks (n== 660, 65.2%) and in Madinah for less than a week (n=614, 60.6%) (Table 2).

### Health Education

The majority of pilgrims had received health education (n=799, 78.9%) and almost a quarter (n=213, 21%) did not receive, and (n=185, 23.2%) selected self-education. The majority received health education in their homeland (n=450, 56.3%), and during arrival (n= 201, 25.2%) the majority believed that the health education they had received was helpful (n=661, 65.3%); however, 13.3% (n=135) disagreed. The most cited source of health education in their opinion were family and friends (n=460, 57.8%), followed by press and publications (n=218, 27.2%), lectures (n=134, 16.8%), and health care providers (n=78, 9.8%), respectively.

### Practice of preventive measures

30.1% of Pilgrims (n=305) never wore masks in crowded areas during Umrah in 2019, in contrast, an almost equal percentage facial masks were always used by pilgrims (n=351, 34.6%) in crowded areas. The majority of pilgrims (n=840, 82.9%) did wash their hands with soap and water or sanitizers after coughing and sneezing. The majority of pilgrims never used an umbrella or hat to protect from the sun, while 144 (14.2%) had (Table 4).

**Table 4.** Responses to items on aspects of practicing of preventive measures by pilgrims (n=1012)

|  | Never<br>F (%) | Rare<br>F (%) | Sometimes<br>F (%) | Often<br>F (%) | Always<br>F (%) |
| --- | --- | --- | --- | --- | --- |
| Using umbrella or hat ☑ | 509(50.2) | 75(7.4) | 130(12.8) | 154(15.2) | 144(14.2) |
| Using sunblock creams ☑ | 652(64.4) | 86(8.5) | 72(7.1) | 79(7.8) | 123(12.1) |
| Drinking at least 8 cups ☑<br>of water | 13(1.3) | 37(3.7) | 151(14.9) | 173(17.1) | 638(63.0) |
| Wearing face masks in ☑<br>crowded areas | 305(30.1) | 7.4(7.3) | 148(14.6) | 134(13.2) | 351(34.6) |
| Sufficient sleep ☑ | 20(2.0) | 45(4.4) | 133(13.1) | 223(22.0) | 591(58.3) |
| Avoid crowded areas ☑ | 70(6.9) | 62(6.1) | 159(19.2) | 207(20.4) | 478(47.2) |
| Using insect repellent spray | 573(56.6) | 107(10.6) | 107(10.6) | 69(6.8) | 156(15.4) |
| Use of personal tools (razor<br>blades, etc.) without sharing with<br>others | 21(2.1) | 4(0.4) | 60(5.9) | 75(7.4) | 852(84.1) |
| Dispose used tissue in waste bin | 18(1.8) | 11(1.1) | 59(5.8) | 100(9.9) | 824(81.3) |
| Washing hands with water, soap or antiseptics |  |  |  |  |  |
| after coughing and ☑<br>sneezing | 3(0.3) | 7(0.7) | 65(6.4) | 97(9.6) | 840(82.9) |
| before eating or ☑<br>preparing food | 3(0.3) | 7(0.7) | 55(5.4) | 87(8.6) | 860(84.9) |
| after using the bathroom ☑ | 3(0.3) | 5(0.5) | 47(4.6) | 90(8.9) | 867(85.6) |

Table 5 displays the responses to items on aspects of availability and the sources of Personal Protective Equipment, where face masks were not available with pilgrims in the majority of responses (n=642, 63.2%).

**Table 5.** Responses to items on aspects of availability and the sources of Personal Protective Equipment (PPE) (n=1012)

|  | Brought it from homeland<br>F (%) | Was given from Al Hamla<br>F (%) | Not available<br>F (%) |
| --- | --- | --- | --- |
| Face masks | 216(21.3) | 154(15.2) | 642(63.2) |
| Personal tools | 230(22.7) | 191(18.9) | 591(58.3) |
| Sanitizers gel | 241(23.8) | 159(15.7) | 611(60.3) |

Table 6 reports the association between receiving health education and practicing of preventive measures by pilgrims, where receiving health education by pilgrims was significantly associated with wearing masks in crowded areas (P= 0.04) and healthy practice score (i.e., washing hands with water, soap, or antiseptics use of personal tools dispose used tissues in waste bin) (P= 0.02).

**Table 6.** Report the association between receiving of health education and practicing of preventive measures by pilgrims

|  | No<br>Median (mean+_SD)<br>Range | Yes<br>Median (mean+_SD)<br>Range | (p) |
| --- | --- | --- | --- |
| Avoiding sun exposure using umbrella or hat | 1(1.9+_1.47) | 2(2.46+_1.6) | 0.03* |
| Using sun block creams | 1(1.45+_1.1) | 1(2.08+_1.51) | 0.01* |
| Drinking at least 8 cups of water | 4.4+_0.9 | 4.35+_0.95 | 0.59 |
| Wearing masks in crowded areas | 1(2.56+_1.77) | 4(3.3+_1.60) | 0.04* |
| Sufficient sleep | 4.74+_0.81 | 4.71+_0.7 | 0.85 |
| Avoid crowded areas | 3.9+_1.4 | 3.97+_1.2 | 0.71 |
| Using insect repellent | 1(2.02+_1.6) | 1(2.17+_1.5) | 0.45 |
| Healthy practice score | 20.5+_4.9<br>7-35 | 22.7+_5.5<br>10-35 | 0.02* |

\* p <0.05 there is a statistically significant difference

Table 7 displays that the majority of pilgrims were very satisfied with the Saudi Arabia Ministry of Health Services (n=734, 72.9%).

**Table 7.** Responses to items on level of satisfaction of Saudi Arabia Ministry of Health Services (n=1012)

|  | F | % |
| --- | --- | --- |
| **The level of satisfaction** |  |  |
| Very satisfied ☐ | 734 | 72.9 |
| Satisfied ☐ | 212 | 20.9 |
| To some what ☐ | 54 | 5.3 |
| Dissatisfied ☐ | 9 | 0.9 |
| Very dissatisfied ☐ | 3 | 0.3 |

## Discussion

This study aimed to assess the experiences of preventive measures practice among 1012 Umrah pilgrims in Saudi Arabia during Umrah season in 2019 (Hijri year 144). 48 different nationalities were linked to the pilgrims who participated in the study survey. The majority of pilgrims in this study had received immunization prior to travel to Saudi Arabia; some had received more than one vaccine, while the most reported immunizations were meningitis, (n=567, 56%), influenza (n=460, 45.5%), and Hepatitis B virus vaccinations (n=324, 32%), respectively (Table 2). However, almost a quarter of the respondents (n=223, 22%) reported not having received any vaccination before travel to the Kingdom.

Notably, all pilgrims arriving Saudi Arabia must have received a single dose of quadrivalent meningococcal vaccine and provide proof of a valid vaccination or prophylaxis 5 years before arrival (13) to allow issuing a Hajj or Umrah visa by the Saudi Arabia Authorities. However, this study reported only 567 (56%) of pilgrims surveyed actually had this vaccination, an issue that should be examined to avoid the spread of this contagious disease (5, 6, 7, 8, 13).

The crowded conditions during Hajj and Umrah increase the risk of respiratory disease transmission (14, 15), including influenzas, tuberculosis, and Middle East Respiratory Syndrome (MERS) coronavirus, which was first identified in Saudi Arabia in 2012 (16, 17, 18). The Centers for Disease Control and Prevention (CDC) strongly recommends that hajjis receive a seasonal influenza vaccine, which is recommended for high risk pilgrims to reduce morbidity and mortality (17). In this study, 45.5% of pilgrims were vaccinated against seasonal influenzas, which is higher than what is recently reported among Malaysian pilgrims (24.9%) (19), French pilgrims (31.8%), and Saudi Arabian, Qatari, and Australian pilgrims (31.0%) (15, 16).

Only 273 pilgrims (26.9%) were vaccinated against polio. The Saudi Arabia health authorities does not require polio vaccine from some countries, but vaccination proof at least 4 weeks before departure is required from pilgrims traveling from polio reporting countries to administer a single dose of the oral polio vaccine. Despite this, free polio vaccination is administered to pilgrims by Saudi Arabian MOH at the entry ports (7).

In this study, the majority of pilgrims had received health education on preventive measures, including aspects of proper hygiene practice (n=799, 78.9%) mostly in their home country (n=450, 56.3%). The current study revealed an association between receiving health education and practice preventive measures by pilgrims, in which receiving health education by pilgrims was significantly associated with wearing masks in crowded areas (P= 0.04) and healthy practices such as washing hands with water, soap, or antiseptics, use of personal tools, and disposal of used tissues in waste bins (P= 0.02).

The majority of respondents believed that the health education they had received was helpful (n=661, 65.3%); however, 13.3% (n=135) disagreed. Considering the experiences of pilgrims with preventive measures, the lowest scores were on use of sunblock creams, insect repellent creams, using umbrellas or hats, and use of face masks in descending order (72.9%, 66.7%, 57.6%, 37.4%). Around 85% of pilgrims always adopted the hygienic practice measures (hand washing and use of personal tools).

According to the pilgrims, different sources were used for health education were typically family and friends (n=460, 57.8%), followed by press and publications (n=218, 27.2%), lectures (n=134, 16.8%), and health care providers (n=78, 9.8%), respectively. However, almost a quarter of the participants reported not receiving any health education and orientation around preventive measures (n=213, 21%). This is obviously a significant issue that added more challenges to the management of the Hajj and Umrah and may have an impact on the organized services that are offered by the Saudi Hajj and Health authorities (7, 10).

Of note, only 485 (47.9%) pilgrims had reported using face masks as “always or often,” and 305 participants (30.1%) had reported never using face masks in crowded areas; however, 63.2% reported lack of availability of these masks. This finding is in accordance with other studies in Saudi Arabia (56%) and the USA (42%) (20, 21).

Since the outbreak of COVID-19 (22), the use of face masks has become universal in China and the rest of the world. However, the evidence of its protection against respiratory infections in the community is still controversial (23). In Saudi Arabia, a study to estimate the incidence of hajj-related acute respiratory tract infection (ARI) among pilgrims travelling from Riyadh revealed that the use of a facemask by men was a significant protective factor against (ARI) in Hajj, and fewer pilgrims (15.0%) had ARI compared with hajjis who used it sometimes (31.4%) or never (61.2%) (24).

It was reported in other studies that the use of face masks for more than eight hours can lead to a substantial decrease in the incidence of influenza-like illness (ILI) among pilgrims (24, 25). However, the effectiveness depended largely on adherence to mask use (26).

In the chain of transmission, regularly washing hands and the use of hand sanitizers causes significantly less ILI symptoms. About 850 of the respondents always performed proper hand hygiene through hand washing during umrah regularly (83.9%). This percentage is higher than the French pilgrims in 2012 (46.3%) and USA pilgrims in 2009 (45.5%) (14, 15).

In the present study, 852 pilgrims (84.1%) always used razor blades for shaving heads, a practice if not hygienic poses a great risk of transmission of blood-borne diseases such as HIV and Hepatitis B and C (5).

Finally, the majority of pilgrims (n=734, 72.9%) rated their experience with the Saudi Arabia Ministry of Health Services as “very satisfied.” Of note is that health services including primary, secondary, and intensive care medical services are offered to millions of Umrah and Hajj pilgrims free of charge.

### Limitations of the study

There are a number of biases that are present in this study, including recruitment by the Al Hamla company members to participate in this study and the responses (the number of refusals to participate was not recorded).

Although the sample size is reasonable and provided detailed and crucial data on the preventive measures, including vaccination history and health education, considering the millions of Umrah pilgrims arrived at the holy cities in Saudi Arabia yearly, low participation and other factors may limit the generalizability of the findings. Time constraints and language barriers were also limiting factors since the recruitment took place at the airport.

Despite these limitations, this study has provided several crucial insights that were consistent with other studies on the Hajj and Umrah in Saudi Arabia.

### Conclusions

This study reported the experiences of 1012 Umrah pilgrims concerning the preventive measures that have been adopted prior and during their religious trip to Saudi Arabia in 2019. The majority had reported taking immunization prior to travel; however, almost a quarter of the respondents reported not having received any vaccinations, including the mandatory meningitis vaccine. The majority of pilgrims had received health education on preventive measures, including hygienic practices, mostly in their home countries. Of note is that almost half of the respondents 485 (47.9%) had reported using face masks “always or often,” however (63.2%) reported that these items were not available. Around 85% of pilgrims always practiced proper hygienic measures (hand washing and use of personal tools).

Preparing awareness messages from this report and strengthening health education would have a beneficial role in shaping personal preventive measures and reducing health hazards.

## Data Availability

The datasets used and/or analysed during the current study are available from the corresponding author on reasonable request.

## Acknowledgments

The authors thank the respondents and their head of campaigns for their participation/cooperation in the survey.

## Study Questionnaire

Dear pilgrim,

The General Administration of Community Participation at the Saudi Ministry of Health and the voluntary Vision Medical team invite you to participate in a research project around the experiences of preventive measures among Umrah pilgrims in 2019 in the Kingdom of Saudi Arabia. Participation in this survey research is entirely voluntary. You can choose not to participate and there are no consequences. You may also withdraw your consent to participate at any time, your participation is completely anonymous and confidential.

**I agree to participate ( )**

**I do not agree to participate ( )**

1. **Your Age?** <20y
  ➢ 20-<35y
  ➢ 35-<50y
  ➢ 50-<60 ?
  ➢ 60 or more
2. **Your nationality** ( )
3. **Your Sex**
  ➢ Male
  ➢ Females
4. **Your Marital status**
  ➢ Widow
  ➢ Single
  ➢ Married
  ➢ Divorced
5. **Your Level of education**
  ➢ Illiterate
  ➢ Read/write
  ➢ Primary /secondary
  ➢ High school or above
6. **The type of chronic disease you have:**
  ➢ Diabetes mellitus
  ➢ Hypertension
  ➢ Cardiac disease
  ➢ Central nervous system
  ➢ Psychological /mental disorders
  ➢ Immunocompromised travelers (sever-moderate)
  ➢ Disabilities (visual-hearing-physical)
  ➢ Others
7. **Your history of vaccination (before travel)**
  ➢ Not vaccinated
  ➢ Influenza vaccine
  ➢ Hepatitis A virus vaccine (HAV)
  ➢ Hepatitis B virus vaccine (HBV)
  ➢ Rabies vaccination
  ➢ Meningitis vaccination
  ➢ Poliomyelitis
  ➢ Pertussis
8. **Duration of stay in Makkah**
  ➢ <1week
  ➢ 1 week-<2week
  ➢ 2week-or more
9. **Duration of stay in Madinah**
  ➢ <1week
  ➢ 1 week-2<week
  ➢ 2week-or more
10. **Did your receive health education on preventive measures to be practiced during Umrah and Hajj**
  ➢ No
  ➢ Yes
  ➢ No, self-education
11. **The site of receiving health education**
  ➢ In homeland
  ➢ During arrival
  ➢ At the residence inside KSA
12. **Did you find the health education that you received beneficial for you to practice appropriate preventive measures during Umrah?**
  ➢ Yes
  ➢ No
  ➢ To some extent
13. **The sources of Health Education**
  ➢ Press and publication
  ➢ Family and friends
  ➢ Lectures
  ➢ Social media (Facebook-Instagram-twitter-snap----------
  ➢ Travel medicine clinics
  ➢ Health care providers
  ➢ Saudi Ministry of Health web site
  ➢ Other web sites
14. **This question is around your actual experiences of the preventive measures you have practiced in the Umrah this year, where Never is the lowest and Always is the highest. Choose the right response for each item.**

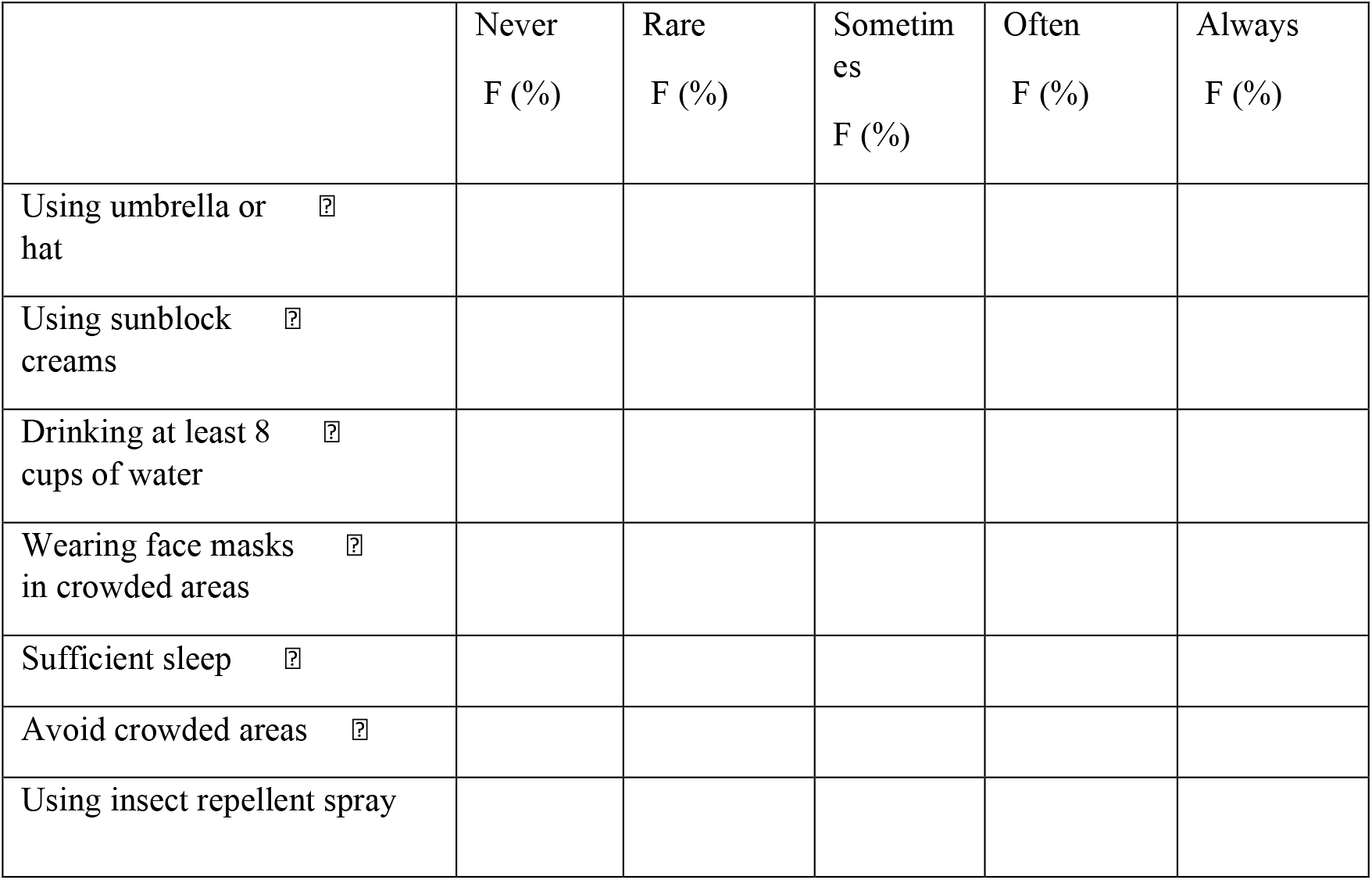
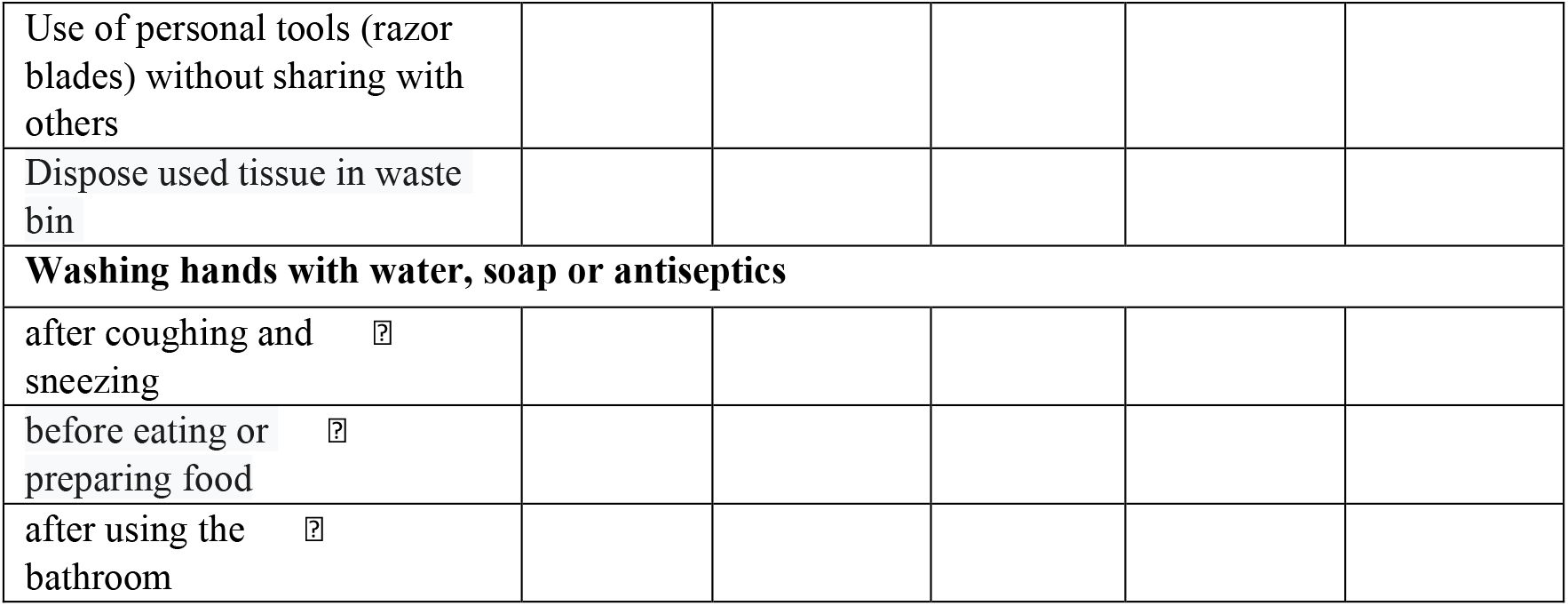
15. **Please can you indicate the availability of the following Personal Protective Equipment (PPE) with you**

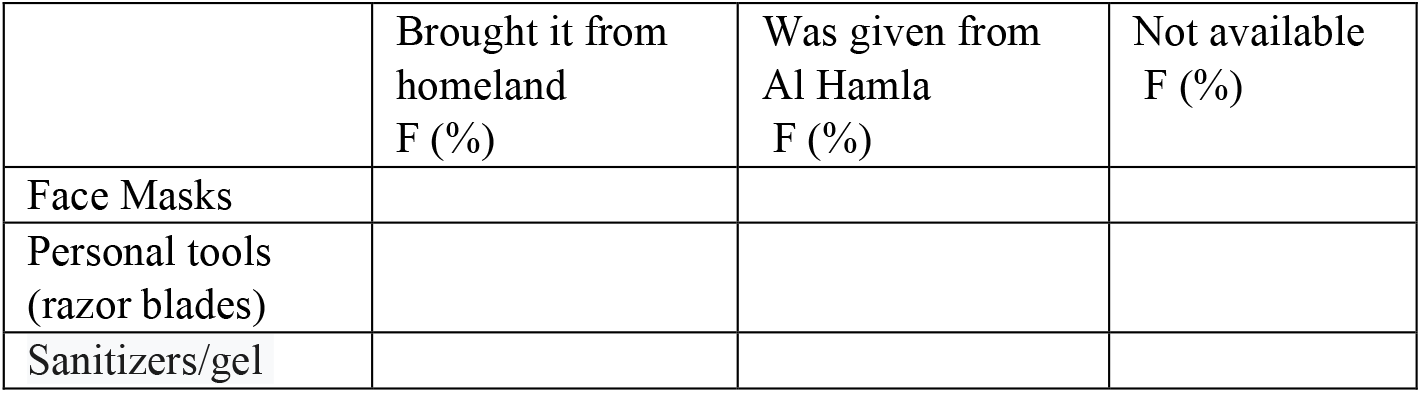
16. **Please can you rate your level of satisfaction of the services offered by ministry of health, including medical care of all types.**
  ➢ Very satisfied
  ➢ Satisfied
  ➢ To some what
  ➢ Dissatisfied
  ➢ Very Dissatisfied

**15- Please can you indicate the availability of the following Personal Protective Equipment (PPE) with you**
|  | Brought it from homeland<br>F (%) | Was given from Al Hamla<br>F (%) | Not available<br>F (%) |
| --- | --- | --- | --- |
| Face Masks |  |  |  |
| Personal tools (razor blades) |  |  |  |
| Sanitizers/gel |  |  |  |

Thank You,

Research Team

